# An approach to quantify parameter uncertainty in early assessment of novel health technologies

**DOI:** 10.1101/2022.02.20.22271248

**Authors:** Rowan Iskandar, Carlo Federici, Cassandra Berns, Carl Rudolf Blankart

**Affiliations:** Center for Evidence Synthesis in Health, Department of Health Services, Policy, & Practice, Brown University, Providence, RI, USA; Center of Excellence in Decision-Analytic Modeling and Health Economics Research, sitem-insel, Bern, Switzerland; Institute of Social and Preventive Medicine, University of Bern, Switzerland; SDA Bocconi school of Management, Centre for Research on Health and Social Care Management (CERGAS), Milan, Italy; KPM Center for Public Management, University of Bern, Bern, Switzerland

**Author notes:** **Corresponding author** Rowan Iskandar, Center of Excellence in Decision-Analytic Modeling and Health Economics Research, sitem-insel, AG, Freiburgstrasse 3, Bern 3010, Switzerland. **Funding statement** This project received funding from the European Union’s Horizon 2020 research and innovation programme under grant agreement 779306 (COMED-Pushing the Boundaries of Cost and Outcome Analysis of Medical Technologies).

**Keywords:** Uncertainty quantification, probabilistic sensitivity analysis, health economic evaluation, cost-effectiveness analysis, early-stage health economic model, probability bound analysis

## Abstract

Health economic modeling of novel technology at the early stages of a product lifecycle has been used to identify technologies that are likely to be cost-effective. Such early assessments are challenging due to the potentially limited amount of data. Modelers typically conduct uncertainty analyses to evaluate their effect on decision-relevant outcomes. Current approaches, however, are limited in their scope of application and imposes an unverifiable assumption, i.e., uncertainty can be precisely represented by a probability distribution. In the absence of reliable data, an approach that uses the fewest number of assumptions is desirable. This study introduces a generalized approach for quantifying parameter uncertainty, i.e., probability bound analysis (PBA), that does not require a precise specification of a probability distribution in the context of early-stage health economic modeling. We introduce the concept of a probability box (p-box) as a measure of uncertainty without necessitating a precise probability distribution. We provide formulas for a p-box given data on summary statistics of a parameter. We describe an approach to propagate p-boxes into a model and provide step-by-step guidance on how to implement PBA. We conduct a case and examine the differences between the status-quo and PBA approaches and their potential implications on decision-making.

## 1 INTRODUCTION

The concept of early-stage health economic modelling has been around since the mid-nineties as a tool to help the decision-making processes of both technology developers and healthcare decision-makers [1-4]. Early-stage economic modelling allows assessing the commercial viability of new technologies in early development phases to guide the development and positioning of the innovation before high investments in further research are made [5]. As an important decision-making tool, an early assessment allows companies to stop further development in the early stages in accordance with the mantra “fail fast, fail cheap” [6]; thereby maximizing the return on investment and societal impact of research and development [7]. For technology developers, the role of early-stage economic models has been explored in several reviews [6, 8-10]. For example, Hartz and colleagues argue that early health economic modelling may support technology developers by providing relevant insights on strategic R&D decision making, pre-clinical preliminary market assessments, go/no-go decisions, development of future trial design, assessment of future reimbursement and pricing scenarios, and price determination [9]. An iterative application from idea screening to market access adds further value to insights of the approach. When iteratively applied, early-stage economic models allow developers to set and revise the envisaged price for the technology as more information on the technology’s performance or on the competitive environment becomes available. In addition, developers may combine early probabilistic models with value of information analyses to guide future research [11]. For healthcare decision-makers, iterative health economic modelling, starting from the early stages of diffusion, has been proposed to precociously identify technologies that are likely to be a good value for money [1] and to balance adoption and reimbursement decisions with the value of further research [12].

However, developers and healthcare decision-makers should remain cautious with the interpretation of the results as the models are informed by data of lower quality and higher uncertainty. In addition, evidence that supports the approach is scarce as comparisons between early- and late-stage models are usually not implemented, although such comparisons would be possible with iterative Bayesian modelling techniques [13]. Grutters et al. [5] demonstrated in their study of 32 applications of early-stage modelling that not a single application resulted in a firm no-go decision. This is because almost all technologies become cost-effective under certain circumstances, e.g., when restricted to a certain subpopulation or to a certain country. Nevertheless, the application of early-stage modelling allows for directing future research in combination with value of information analyses, readjusting pricing expectations [11], and initiating an early dialogue between the developers of new technologies and pricing and reimbursement agencies [14].

Similar to later stage models, correct characterization of the uncertainty around model parameters and how it propagates to the outputs of interest (e.g., the technology’s cost, effectiveness, or cost-effectiveness) is a key component of the modelling approach [15]. At its most basic characterization, parameter uncertainty means that we do not know the exact value of a parameter for reasons such as finite sample size, low data quality, and different study settings or populations (non-transferability of estimates) [16]. In the context where data is available, we can leverage standard statistical techniques to represent uncertainty in the form of a probability distribution. However, when assessing the cost-effectiveness of a novel intervention early in its life cycle, data and knowledge are typically limited or even non-existent. Previous early models have addressed this issue by conducting univariate or bivariate sensitivity analyses on individual parameters [5, 6]. Although deterministic sensitivity analyses provide some indications about the importance of individual parameters, they do not provide any information on the uncertainty in the outputs of interest. The ISPOR-SMDM best-practice [17] recommends only two analytical tools for modelling parameter uncertainty. First, probabilistic sensitivity analysis (PSA) is recommended as a full characterization of the uncertainty around model’s parameters by defining a set of default probability distributions that are often parametric. The choice of the parametric distribution is mainly driven by the consideration of the parameter’s support. For example, a beta distribution is used for characterizing the uncertainty of a parameter with a support [0, 1]. Forcing the modelers to commit to a particular distribution implicitly assumes that the modelers have more information (e.g., knowing the shape of a distribution) than they actually possess, and the uncertainty is known and quantifiable by a probability distribution [18].

However, when conducting early economic models, we may have only partial or no information about the probability distribution, i.e., we cannot assign the relative plausibilities of different parameter values, and therefore, conducting PSA may be impracticable or even lead to misleading conclusions [5]. When no prior data is available to define parameters’ probability distributions, the best practice proposes the use of expert knowledge elicitation [18]. However, for technologies in their early phase of development, experts may not have yet formed a belief over the performance of the new technology, making the elicitation process more challenging. For example, for very innovative medical devices, experts may not have an idea of how the new device will affect patients’ outcomes or reduce resource consumption and costs until they gain some experience with using it. To handle such data sparsity situations, it is desirable to have an approach for quantifying parameter uncertainty without the need for assuming precise probability distributions.

Iskandar [19] recently introduced the probability bound analysis (PBA) approach to quantify parameter uncertainty where the only information accessible to practitioners include the combinations of summary statistics (mean, median, and standard deviations), quantiles, and minimum and maximum of a parameter (hereafter collectively termed as *minimal data*). In PBA, the uncertainty about the probability distribution for each parameter is expressed in terms of upper and lower bounds on the cumulative distribution function (CDF). The area bounded by these upper and lower bounds is guaranteed to contain the unknown CDF and can be derived from minimal data. Prior to our introduction to the concept, the idea of bounding unknown probabilities given minimal data was primarily employed in risk analysis [20] and relatively unknown to the fields of health economic evaluation. All known approaches for estimating bounds on the unknown CDF, e.g., Kolmogorov–Smirnov limits [21], require datasets. Furthermore, to date, no study has explored the application of PBA specifically in early-stage health economic modelling where the problem of data sparsity is prevalent.

Building upon the methodological work [19], the aim of the paper is two-fold. First, we describe an application of PBA for quantifying parameter uncertainty in the context of minimal data that characterize early-stage health economic models. Second, we provide step-by-step guidance for implementing PBA in a generic mathematical model. For this study, we assume that the model parameters are mutually independent [22]. This paper is organized in the following way. First, we review the concept of parameter uncertainty quantification and the status-quo (PSA) approach. Secondly, we formally introduce PBA and provide relevant formulas. This study focuses on *free probability boxes* that are a generalization of parametric probability boxes. Then, we introduce an approach for propagating parameter uncertainty into a mathematical model. Next, we provide guidance on how to conduct PBA. Lastly, we demonstrate an application of PBA in health economic modelling of a hypothetical new total artificial heart for the treatment of patients with advanced biventricular heart failure.

## 2 PARAMETER UNCERTAINTY QUANTIFICATION

This section starts with a summary of the concept of parameter uncertainty quantification and the current approach, probabilistic sensitivity analyses (PSA), and ends with an introduction to the concept of PBA as an alternative approach.

### 2.1 Concept

We consider a health economic model ℳ that is parameterized by a set of model parameters, *θ* where the true values of *θ* are unknown due to lack of data (*epistemic uncertainty*). We want to estimate the consequence of uncertainty in the values of the parameters on decision-relevant outcomes *Y* (*parameter uncertainty quantification*) by conducting the following tasks. First, we prescribe a theoretical framework to represent the varying degrees of uncertainty in the model parameters (*parameter uncertainty representation*). Then, we provide an approach for propagating parameter uncertainty into our health economic model (parameter uncertainty propagation). This task will generate measures of uncertainty in outcomes. Lastly, we specify ways to utilize the uncertainty in the model outcomes for use in further analytical tasks.

### 2.2 Probabilistic Sensitivity Analysis

Following the above steps, the status-quo approach for parameter uncertainty quantification, i.e., PSA [17], starts by treating the model parameters or their sample means as random variables that are endowed with probability distributions. If the sample size of the data informing the parameter is large enough, then by the central limit theorem, the sampling distribution of the mean will converge to a normal distribution, which then can be used in PSA [17]. However, when the sample size is small, using normal distribution could lead to unplausible values. Therefore, it is a common practice to select a type of probability distribution whose support matches with the model parameter’s domain (e.g., beta distribution for probability parameters with domains [0,1]). The location and ancillary parameters of the chosen distributions are then estimated using either individual-level data or a moment matching approach from aggregated secondary data [15]. After we have assigned probability distributions to every uncertain parameter, we typically use an iterative Monte Carlo sampling approach [23] for the parameter uncertainty propagation task. We sample parameters values independently from their probability distributions and evaluate the model at each sampled set of values. Following the propagation task and given the resulting empirical distributions of the model outcomes, we can estimate their expected values and use them as inputs for calculating the cost-effectiveness ratios and other measures used for decision making.

### 2.3 Probability Bound Analysis

The starting point of PBA is akin to that of PSA, i.e., model parameters are treated as random variables. However, PBA imposes a weaker restriction, i.e., the relative plausibility of parameter values cannot be defined in terms of precise probability distributions. PBA treats the cumulative distribution function (CDF) of a parameter as unknown and provides lower and upper bounds to the CDF instead, which are, termed as lower-bounding function (LBF)*<u>F</u>*(*θ*), and an upper-bounding function (UBF) 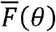, respectively. We define a *p-box* as a set of CDFs of a parameter θ (F(θ), i.e., 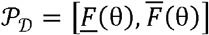, where each CDF belonging to the set is consistent with a given minimal data 𝒟.[24, 25] For each realization of *θ*, e.g., 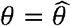, the epistemic uncertainty is represented by the interval 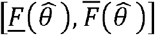 In other words, we can only say that the value of 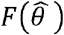 is known up to an interval due to our lack of knowledge about the precise form of the CDF. As a corollary, the more information about the CDF we have, the tighter the area enclosing the unknown CDF becomes (Figure 1). In fact, if the CDF is fully known, the interval degenerates to a single point, i.e., the p-box coincides with the CDF. Based on this observation, the p-box is dependent on the available data. We derive p-box formulas for different subsets of minimal data and provide them in Appendix A.1. For details on the derivations of the formulas, interested readers should consult [26]. After specifying the p-box for each parameter, we propagate the parameter uncertainties represented by the p-boxes into our health economic model, which is described in the next section, to generate a p-box of the model outcome, i.e., 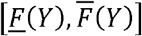.

**Figure 1.**
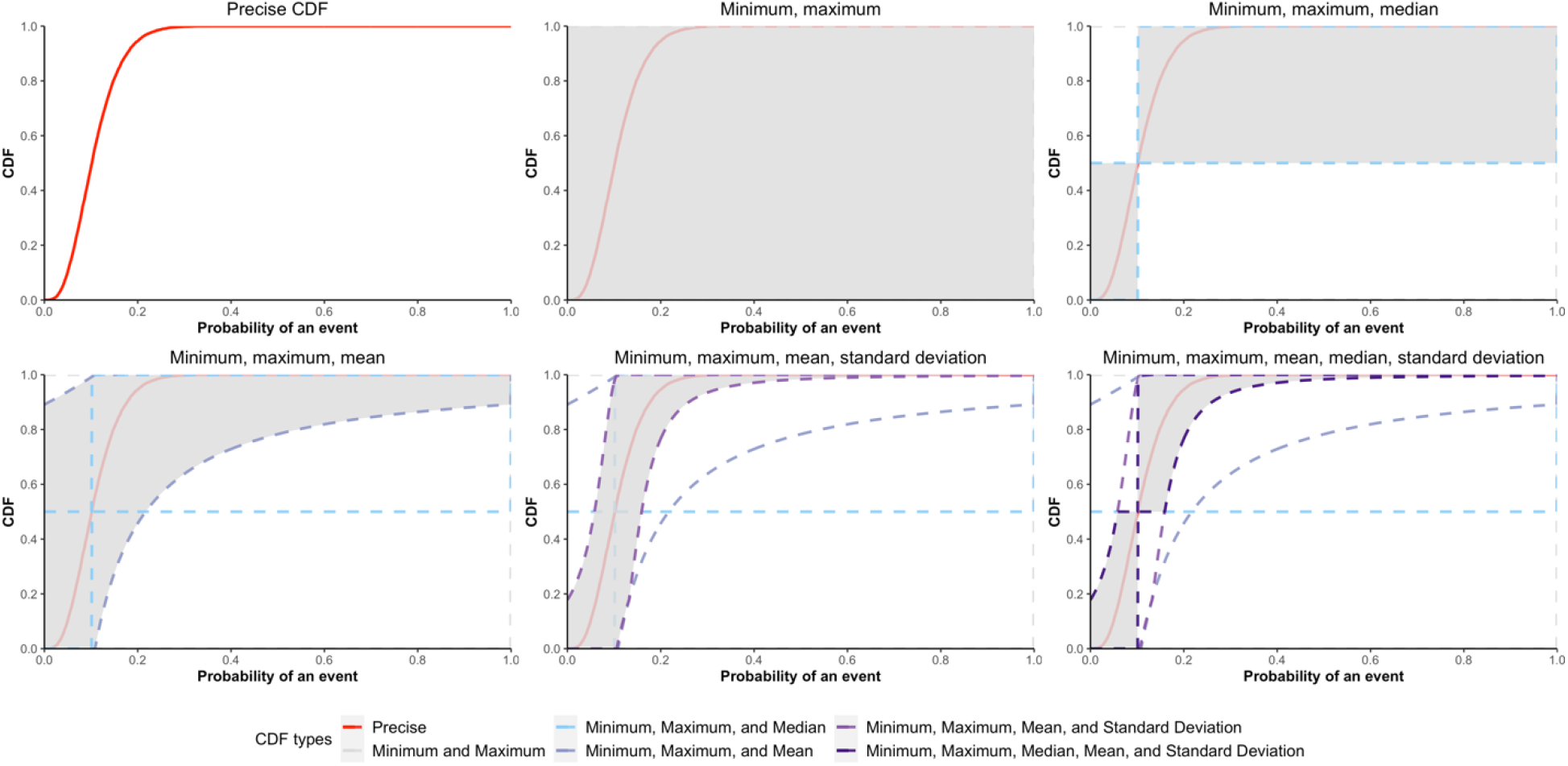
P-boxes for a generic probability of an event under different minimal data and the true unknown cumulative distribution function of the parameter. The shaded region is the area between the lower bounding function and the upper bounding function for a given minimal data. As we have more data, the region shrinks. CDF: Cumulative distribution function.

For the interpretation and utility of having the uncertainty in outcomes represented by a p-box, we note that the calculation of the expected values of model outcome *Y* over its p-box results in an interval of expected values, i.e.,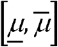, where 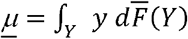 and 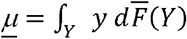. The interval includes all expected values that correspond to CDFs that are enclosed by the p-box. This is true because the p-box of *Y* is guaranteed to enclose all CDFs of *Y*. Given what we know and assume about the uncertainty of the model parameters, the expected outcomes cannot be larger (smaller) than the expected value of *Y* over its upper (lower) interval bound. In contrast to assuming precise CDFs for all parameters where the expected value of a decision-relevant outcome is a single value (vs. an interval of values), alternative decision rules are needed as expected value maximization is no longer amenable. One compatible decision rule is the Hurwicz decision criterion, where the lower and upper expected values correspond to the best- and worst-case scenarios, respectively. The Hurwicz method is typically used to determine the value of an intervention by factoring in both the worst- and best-case scenarios. This value is estimated by choosing an α value, which represents how optimistic a decision-maker is about the best-case scenario and calculating a weighted average of the best-case and worst-case outcomes, which are represented by 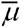 and *<u>µ</u>*, respectively. More formally, given an α, the value of an intervention is equal to 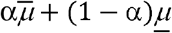. The optimal intervention is the one that has the highest values among the alternatives.

## 3 IMPLEMENTING PBA

This section describes the steps for implementing PBA in a health economic model and closely follows the steps outlined in [19]. We start by briefly reviewing parameter uncertainty quantification using PSA and drawing upon the similarities between PBA and PSA to build intuitions of how to implement PBA in practice.

The uncertainty propagation in the PSA context works by sampling a parameter value from its precise CDF. The sampling can done by using the inverse transform sampling method if the inverse of the CDF is explicitly known. In PBA, the uncertainty propagation also uses sampling, albeit with one key difference, i.e., we sample an interval of values instead of a single value. The sampling procedure follows the idea of an inverse transform sampling. For each value in the image of the LBF and UBF of a parameter (in the probability space [0,1]), we sample an interval of the values (in the domain of the parameter) by using the inverses of the LBF and UBF. Instead of sampling all possible disjoint intervals for all values in [0,1], we consider coarser discretization of the image of the LBF and UBF into *sub-intervals*. The coarser the discretization is (fewer sub-intervals), the less accurate the uncertainty propagation becomes. For each sub-interval and its endpoints, we calculate the corresponding interval of parameter values using the inverse of the p-box. The choice of how to evaluate the endpoints of the sub-interval, e.g., use the inverse of the LBF (UBF) for the upper (lower) endpoint, determines the accuracy of the approximation due to discretization. The probability of observing a particular sub-interval of the parameter value is equal to the length of the sub-interval in the probability space. The sampling of the sub-intervals and the calculation of their probabilities are then repeated for each parameter. Since there are multiple possible realizations for each parameter, which is equal to the number of sub-intervals, we must consider all possible combinations of sub-intervals across all parameters. Because of our assumption about the independence among parameters, we compute the probability of each possible combination (henceforward termed as a hyperrectangle) by multiplying the probabilities assigned to the sub-intervals comprising the combination. After specifying an approach for sampling hyperrectangles from p-boxes, we must prescribe a method to evaluate our model at each sampled hyperrectangle. One intuitive approach is based on optimization, where we find a pair of optima, i.e., the minimum and maximum values of the model outcome for each hyperrectangle. We set the probability of observing a pair of optimum values equal to the sampling probability of the corresponding hyperrectangle. To estimate the p-box of the model outcome empirically, we cumulate the probabilities of each minimum (maximum) to derive the UBF (LBF). The parameter uncertainty quantification procedure for PBA is summarized in the following steps.

**S1**: We decide which model parameters are suitable for PBA, comprising the set *θ*_*p*._

**S2**: For each PBA parameter in *θ*_*p*_ (*θ*_*i*_ *∈ θ*_*p*_) we determine the available minimal data 𝒟_*i*_ where *i* indexes the model parameter.

**S3:** For each *θ*_*i*_, we partition the interval [0,1] (the probability space) into a finite number of intervals (sub-intervals), where each parameter may have a different number of sub-intervals, *n*_*i*._ The finer the discretization is (higher *n*_*i*_), the higher the accuracy of the approximation of the p-box of *θ*_*i*_ (Figure 2) and the p-box of the model outcome will be. The optimal choice of *n*_*i*_ is problem-dependent and can be determined in the following way. First, we choose a decision-relevant metric whose error we aim to minimize and set an error tolerance and a computational budget in terms of computational time. Next, we choose a benchmark *n*_*i*_, which should be a relatively high number. As a measure for error, we calculate the relative difference in the average decision-relevant metrics between those obtained from running PBA (steps S4 to S7) at the benchmark and a particular *n*_*i*_. Then, we start with a low number of *n*_*i*_ and increment the number only if the following is true: the resulting error or the computational time are less than or equal to the error tolerance and maximum computational time, respectively. After finding the optimal *n*_*i*_, we assign a weight to each sub-interval 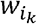, where *i*_*k*_ indexes the *k*-th sub-interval of *θ*_*i*_ A typical weight is a uniform weight for all sub-intervals, i.e., 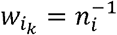.

**Figure 2.**
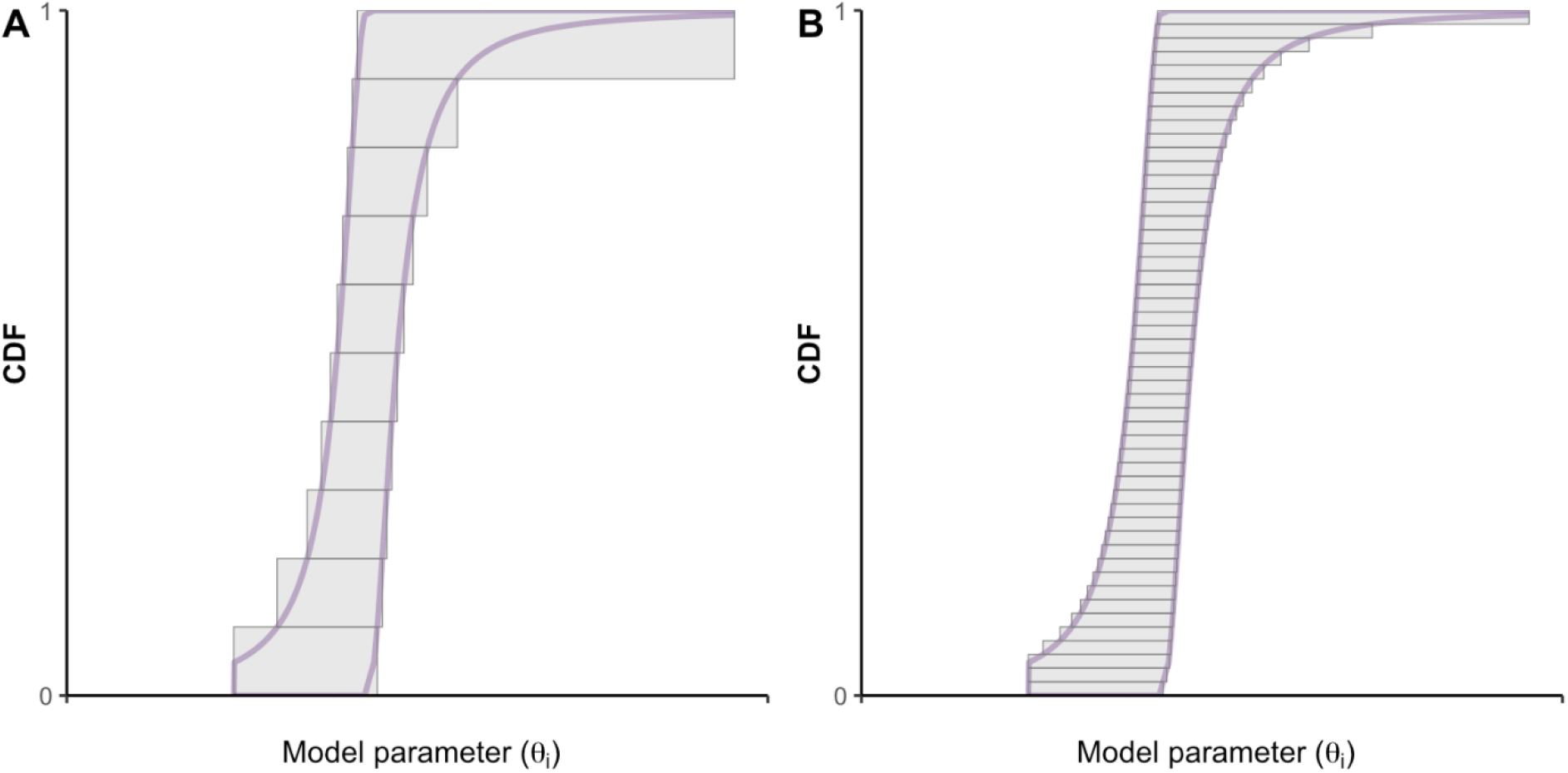
Approximation of a p-box (purple lines) using the outer discretization approach (grey boxes). Sub-figures A and B show the accuracy of the approximation when using, =10 and =50, respectively. CDF: Cumulative distribution function.

**S4:** For each sub-interval of a parameter, we determine the lower and upper endpoints and calculate their corresponding upper and lower endpoints in the parameter space using the quasi-inverse of the LBF and UBF, conditional on 𝒟_*i*_. One approach to determine these endpoints is based on the outer discretization method (Figure 2) [26]. This step generates the sub-intervals in the parameter space.

**S5:** Given the sub-intervals, we enumerate all possible combinations, where each combination is formed by intersecting a particular set of sub-intervals of all parameters. The number of combinations is determined by the number of sub-intervals of each parameter.

**S6:** For each combination of sub-intervals, we compute the minimum and maximum of the model outcome using an optimization method over the parameter space defined by the combination. This step produces intervals constructed from the minimum and maximum values of *Y* and their associated weights. The weight for each interval of *Y* is equal to the weight of the corresponding combination of sub-intervals. The choice of the optimization method is problem-dependent and typically determined by, for example, the model characteristics such as non-linearity and convexity of the model, the availability of information on the gradient, or the trade-off between computational budget and accuracy.

**S7:** To derive the p-box of, we calculate two empirical CDFs by cumulating the weights for the values of the minimum and the maximum are less than some arbitrary values in the support of

In some cases, we may have access to information that is sufficient for specifying probability distributions of some parameters, comprising the set To propagate uncertainty from both parameter sets and into we include the following steps.

**S0:** We conduct *N* repeated samplings from the precise CDFs of parameters in to generate a sequence of *N* samples. For each sample of, we repeat steps **S2** to **S6**.

**S8**: The average p-box of is calculated by averaging p-boxes over the *N* samples.

## 4 APPLICATION OF PBA

We conduct a case study that exemplifies an early assessment of novel health technology to demonstrate how to implement PBA and the difference between PBA and PSA.

### 4.1 Economic model of a novel total artificial heart

Heart transplantation is the optimal treatment for patients with advanced heart failure (HF) [27, 28]. However, mainly due to limited donors’ heart availability, The use of ventricular assist devices to bridge patients until a new heart becomes available is often necessary. For patients with particularly severe conditions, completely substituting the sick heart with a total artificial heart (TAH) has proven to be beneficial [27]. Until recently, only Syncardia TAH was approved as a bridge-to-transplant therapy, but other more advanced models are under development or recently got regulatory approval [27]. Evidence on ventricular assist devices is generally poor at the early stages of adoption, with very few randomized clinical studies comparing the devices and therapeutic strategies [29] [30] [28]. Indeed, devices usually gain market access despite limited evidence on their relative (cost-)effectiveness, and their performance is then monitored retrospectively through multinational registries such as the EUROMACS registry[31] or the INTERMACS [32]. Consequently, developing health economic models to explore the technology’s cost-effectiveness can be particularly challenging and require strong assumptions on model inputs [33].

To demonstrate the utility of PBA in this setting, we used a simplified version of a previous cost-effectiveness model of a hypothetical new TAH versus Syncardia. Full details of the model are provided elsewhere in this supplement [34], and summarised here. The model was structured as a discrete-time semi-Markov model with three states, alive with TAH support (TAH), alive after transplantation (HTx), and dead (Figure 3).

**Figure 3.**
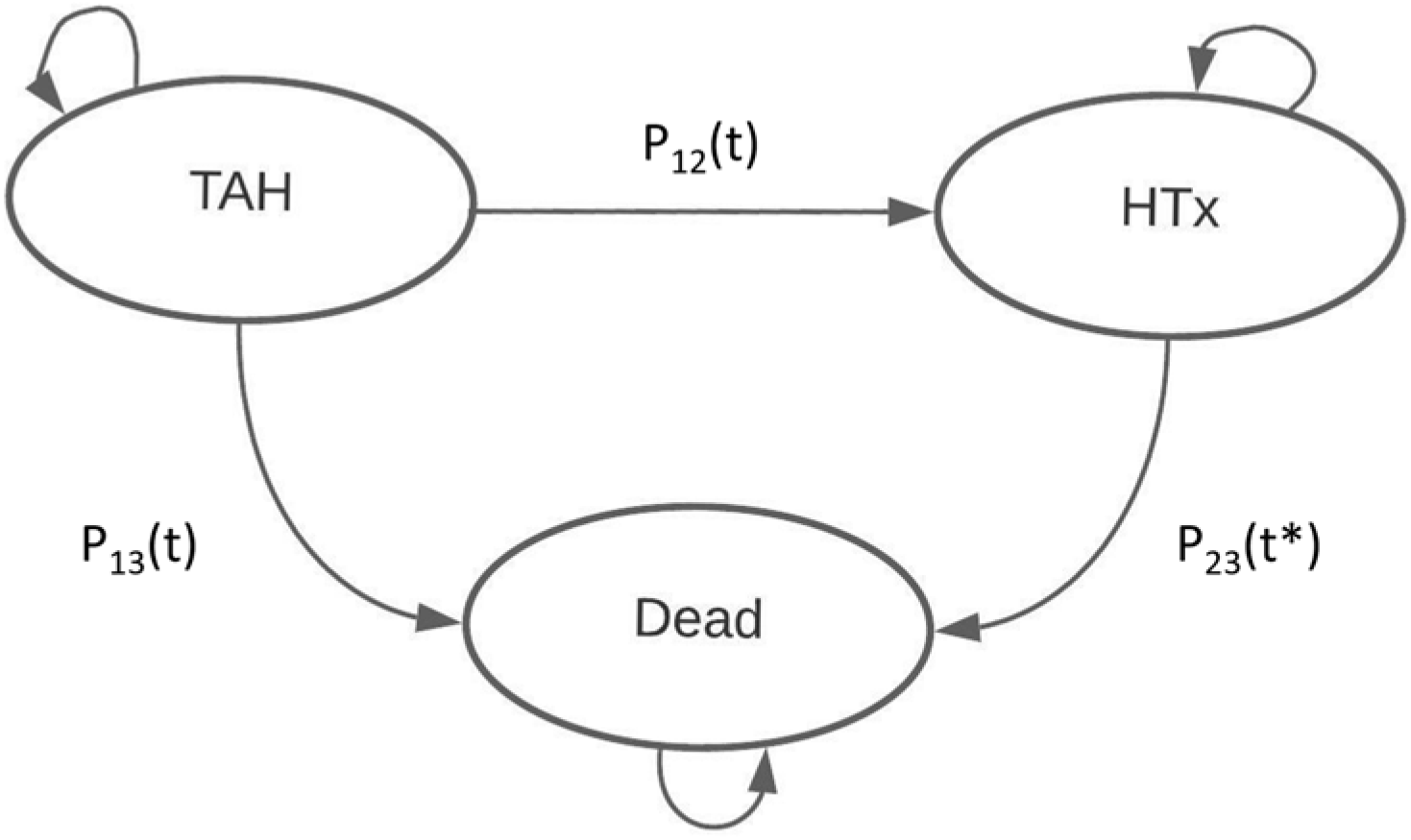
Graphical representation of the TAH Model. *The transition probabilities from the TAH to the HTx state depends on the time of entrance in the state. TAH: Total artificial heart, HTx: heart transplantation.

Probabilities of death from either the TAH or HTx state were modelled as a function of time since the time of entrance in that state. Following Sharpless et. al. [35], the model was decomposed into two simple sub-processes, one representing time pre-transplantation and the second representing time post-transplantation. During support with HTA, the probabilities of having either HTx or dying were modelled as a simple competing risk model attributing either the HTx or dead state conditional on leaving the TAH state. Data for the Syncardia was retrieved from secondary sources from the literature, including survival data from prospective cohort studies, previous cost-effectiveness studies on other MCS devices, and other secondary data on costs and utilities (Appendix A, Table A.2). For the novel TAH, data from a single-arm pivotal study with 35 patients and 24 months follow-up was simulated. We arbitrarily assumed that patients would have similar times on support before experiencing either death or HTx, but that the risk ratio of death between the novel and Syncardia at each time point was lower by 25% due to the higher chance with the new device of surviving until a donor’s heart became available. Similarly, a 15% reduction in the probability of any major adverse event was simulated. The changes in the rate of adverse events was linked to changes in costs, but not in mortality or utilities due to the lack of information on the correlation structure between adverse events and these outcomes. A long-term utility decrement was only applied to patients who experienced disabling strokes. The simulated parameters for the novel TAH are reported in Table A.3 (Appendix A). Procedure costs were considered equal to the Syncardia except for the cost of the device. During support, monthly differences in costs were originated from the model based on differences in the number of discharged patients and the incidence rates of adverse events. Patients surviving to HTx were assumed to have the same monthly costs as the two devices. The cost of the novel TAH was arbitrarily set at €200,000. Cost-effectiveness was estimated by calculating incremental net-monetary benefits (INMBs) using an arbitrary cost-effectiveness threshold of €30,000 per QALY gained. The model is coded in R [36] and is available under a GNU GPL license and can be found at https://github.com/rowaniskandar/TAH-PBA.

### 4.2 Uncertainty Quantification

We quantify the effect of uncertainty in some parameters on the incremental net monetary benefit (INMB) and follow the steps outlined in Section 3.

**S1:** We choose model parameters *θ*_*i*_ that are characterized by lack of data for the novel TAH: (1) probability of dying in the first month, (2) probability of dying in the second month, (3) probability of dying in the third month, and (4) probability of major device malfunction. These four parameters comprise the set *θ*_*p*_. Note that the same parameters have been used for PSA to allow comparison between the two approaches, whereas all other parameters in the model have been considered fixed. Table 1 reports the summary statistics and assumptions used in PBA and PSA.

**Table 1.** Parameters used for the probabilistic sensitivity analysis and the probabilistic bound analysis.

| Model parameters | Parametric distribution used in PSA | Data used in PBA and PSA |  |  |  |  |
| --- | --- | --- | --- | --- | --- | --- |
|  |  | Minimum | Maximum | Median | Mean | Standard deviation |
| Probability of death<sup>1</sup> |  |  |  |  |  |  |
| 1 month | Beta | 0 | 1 | 0.62 | 0.61 | 0.13 |
| 2 month | Beta | 0 | 1 | 0.1 | 0.12 | 0.11 |
| 3 month | Beta | 0 | 1 | 0.11 | 0.14 | 0.12 |
| Probability of major device malfunction | Beta | 0 | 1 | 0.1 | 0.11 | 0.05 |

<sup>1</sup> while on support conditional on leaving the TAH state
PBA: Probability bound analysis, PSA: Probabilistic sensitivity analyses

**S2:** For each parameter in *θ*_*p*_, we assume to have data on the minimum, maximum, mean, median, and standard deviation, comprising 𝒟_*i*._

**S3:** For each *θ*_*i*_, we partition the interval [0,1] into 35 sub-intervals(*n*_*i*_ =35) and assign a uniform weight to each sub-interval 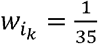. We set an error tolerance of 1% with INMB as the metric and a maximum computational time of 12 hours. We used *n*_*i*_ =50 as the benchmark and tested *n*_*i*_s sequentially starting from five in increments of five. We determined 35 as the optimal number of sub-intervals that satisfies both conditions (an error of 0.33% and a computational time of 11.2 hours).

**S4:** For each sub-interval of *θ*_*i*_, we calculate their corresponding upper and lower endpoints in the parameter space, using the quasi-inverses. The quasi-inverses of the p-box constructed from data on the minimum, maximum, mean, median, and standard deviation are derived from two sets of quasi-inverses, i.e., quasi-inverses of p-box given minimum, maximum, and median (Equations A.7 and A8), and quasi-inverses of p-box given minimum, maximum, mean, and standard deviation (Equations A.15 and A.16). We then intersect (using Equations A.17 and A.18) the two quasi-inverses of the LBFs and UBFs to derive the desired p-box of *θ*_*i*_.

**S5:** Given the sub-intervals, we enumerate all possible combinations by intersecting a particular sub-interval for each parameter, giving a total of 15^4^ combinations.

**S6:** To compute the minimum and maximum of the INMB, we apply a deterministic search algorithm based on systematic divisions of the domain (each combination of sub-intervals) into smaller hyperrectangles [37] and use the implementation of the *nplot* library [38] in the R program.[39]

**S7:** To derive the p-box of the INMB, we calculate two empirical CDFs by cumulating the weights for the values of the minimum and the maximum that are less than some arbitrary values in the domain of *INMB*, from -200,000 to 125,000.

To demonstrate the difference between PSA and PBA, we conduct a PSA using beta distributions with the same summary statistics to represent uncertainty in the four parameters exemplifying the common practice of using “off-the-shelf” distributions given minimal data. To show how the two methods differ in encoding our complete lack of data, i.e., knowing only the minimum and maximum values of the four parameters, we use uniform distributions and compare the status-quo approach with PBA. We then demonstrate how to conduct decision analysis using the results of PBA.

### 4.3 Interpretation of Results

The first comparison shows the difference between the results of parameter uncertainty quantification using precise CDFs vs. PBA. The INMB shows the difference in net monetary benefit between two alternative health technologies, i.e., the hypothetical TAH compared with the Syncardia. A positive INMB indicates that the hypothetical new TAH is cost-effective compared to the Syncardia, given a willingness-to-pay threshold of €30,000 per QALY. For a given minimal data, PBA results in a p-box enclosing the unknown CDF of the INMB (Figure 4). In contrast, by imposing additional parametric assumptions on parameter distributions (beta distributions), PSA yields an empirical CDF of the INMB, resulting in a mean INMB of €32,588 (95% credible interval of [€1,313, €63,865]) (Figure 4). When compared to PSA, PBA gives two additional information. First, PBA provides information on the amount of uncertainty in the INMB due to our imperfect or complete lack of knowledge about some parameters, which is indicated by the area enclosed by the p-box. Secondly, PBA provides bounds on the plausible values of the INMB. The minimum is given by the minimum of the INMB values where the UBF assume a non-zero value (-€172,383) and the maximum is given by the minimum of the INMB values where the LBF value is equal to one (€115,242). Simply put, the true INMB value lies in the interval that is bounded by these two extreme values. We also observe that the area bounded by the corresponding UBF and LBF shrinks as we have more data (Figure 4). This is expected; as more data on the CDF becomes available, we can pinpoint the location and shape of the CDF more accurately. Although the p-box becomes smaller with more information, the area still extends to the left of zero. Since we have no information about the uncertainty within the p-box of the INMB, we cannot make any definite conclusions about the probability of whether the hypothetical TAH is more cost-effective compared to Syncardia. Nevertheless, PBA is transparent about the possible range of cost-effectiveness outcomes, i.e., our approach does not restrict the range artificially by making assumptions that are not supported by data.

**Figure 4.**
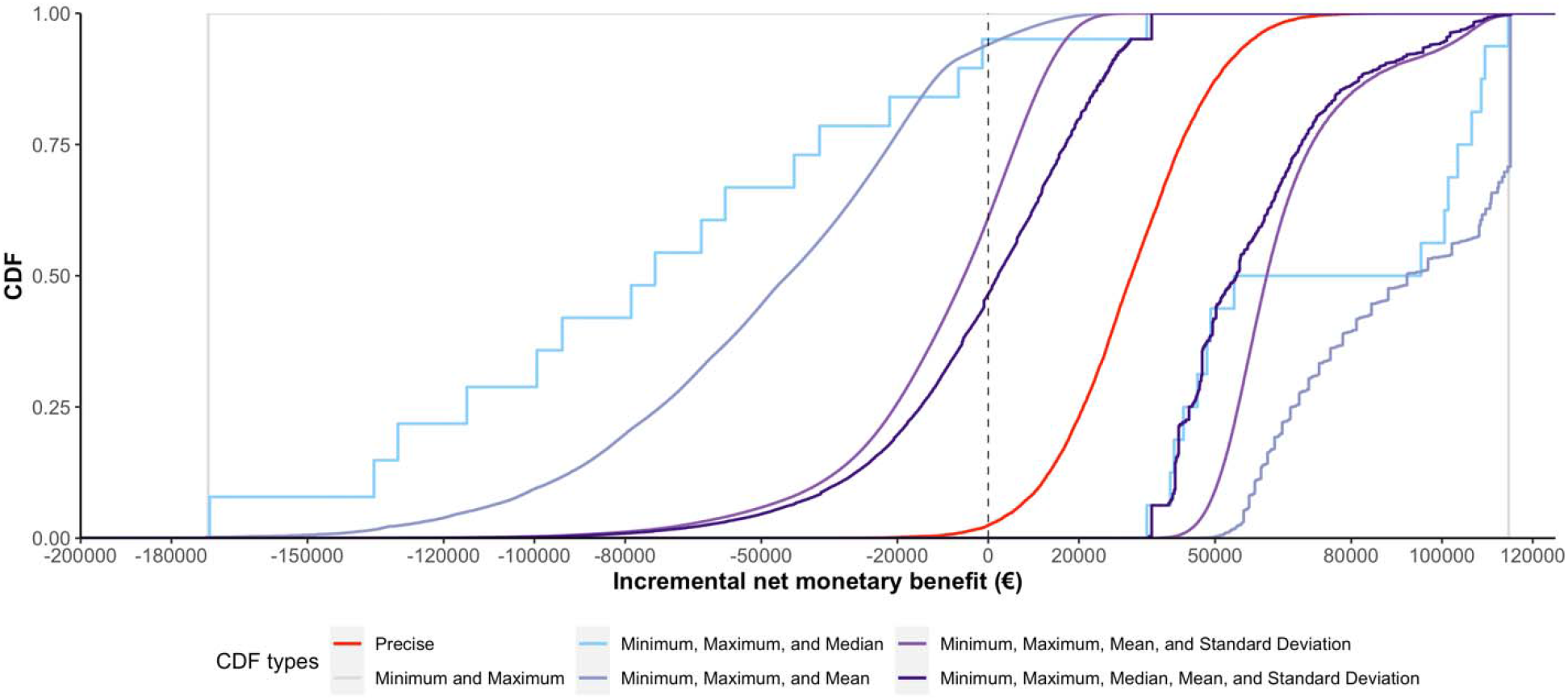
Uncertainty around incremental net monetary benefits of novel TAH using p-boxes under different minimal data vs. beta distributions for four parameters. CDF: Cumulative distribution function.

The second comparison demonstrates the consequence of assuming more information than we actually have. Assuming each value in the interval [0, 1] is equally likely is *qualitatively* and *quantitatively* different from knowing that the value is somewhere in the interval. A PBA does not provide any information on the uncertainty in the INMB, given that the uncertainties in the parameters are represented by intervals with no measures of uncertainty assigned to the intervals. In other words, a PBA transfers our ignorance in the model parameters to ignorance in the model outcomes. In contrast to PBA, a PSA using uniform distributions does not preserve our ignorance, i.e., uniform distributions in the model parameters do not generally translate to a uniform distribution in the model outcomes unless the model is linear [40]. Furthermore, using uniform distributions generates a “precise” CDF (Figure 5), hence creating a pseudo-certainty in the model outcome. In sum, PBA uses fewer assumptions and is more conservative in its estimates of the resulting uncertainty. Such conservativeness may be desirable if an erroneous decision will lead to a catastrophic loss.

**Figure 5.**
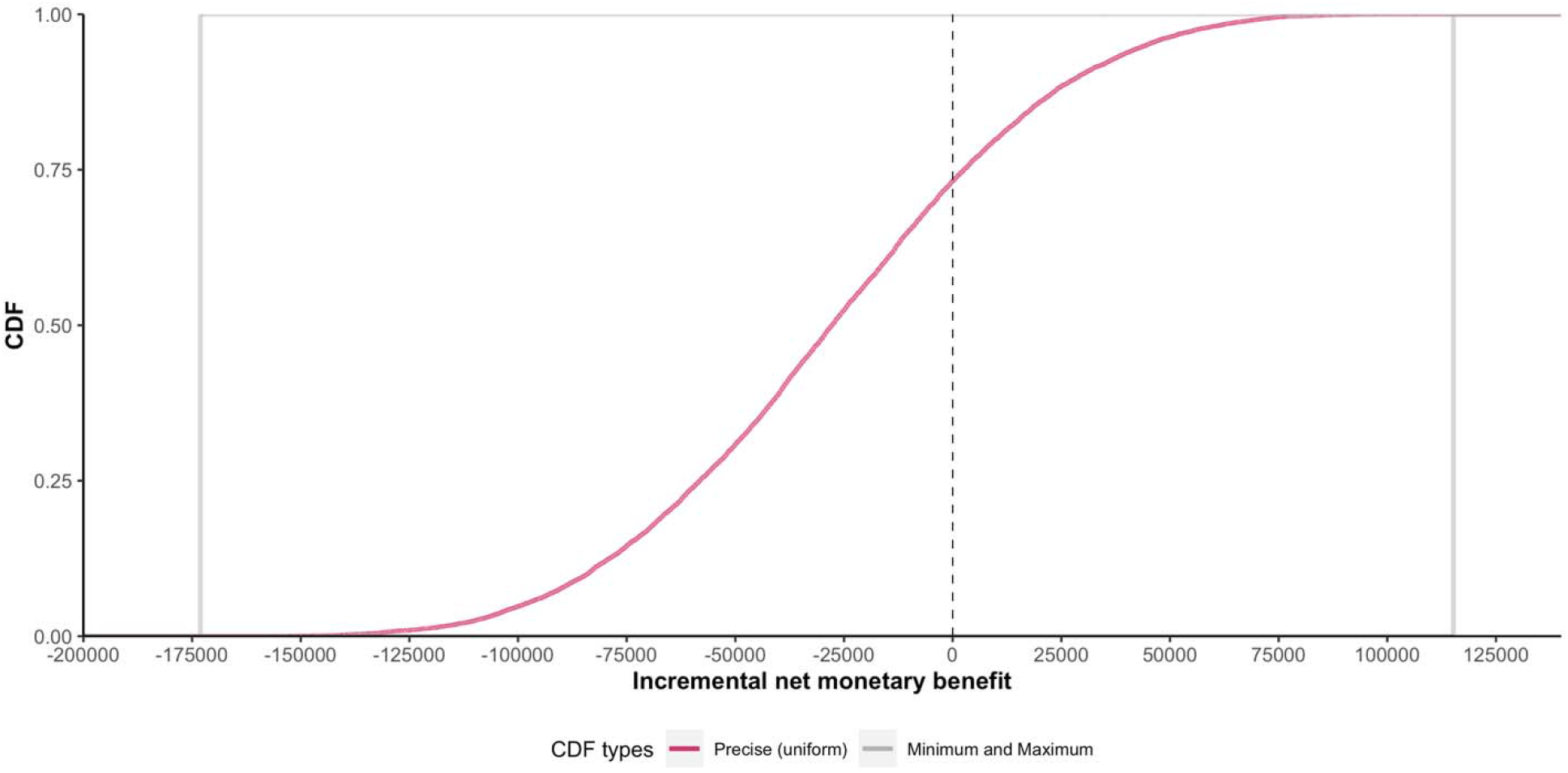
Uncertainty around incremental net monetary benefit of TAH model using p-boxes given minimum and maximum values vs. uniform distributions using the same minimum and maximum values for the four parameters. CDF: Cumulative distribution function.

To determine whether the novel TAH is cost-effective based on the PBA results, we use Hurwicz’s criterion. For each, we calculate _and _ by averaging UBF and LBF over INMB values, respectively. Then, for each α, we compute the weighted average and determine whether the INMB is greater than 0 (cost-effective). Table 2 shows the results using several representative α values. We first note that the interval of the expected INMBs, i.e., _ _shrinks as we have more data. As the decision-maker becomes less optimistic, i.e., lower α, the novel TAH has lower INMBs for a given. For example, if the available data consist of only minimum, maximum, and mean values (or fewer), then the novel TAH is not cost-effective at α=0.25 or lower. Conversely, if the decision-maker’s attitude is represented by a particular α, then the novel TAH becomes more likely to be cost-effective as we have more data. In the extreme situation where the decision-maker is fully confident that the best-case scenario will occur, the amount of data has no influence on the cost-effectiveness of the novel TAH. Similarly, when the decision-maker is fully pessimistic, the novel TAH is deemed to be not cost-effective regardless of how much data we have.

**Table 2.**
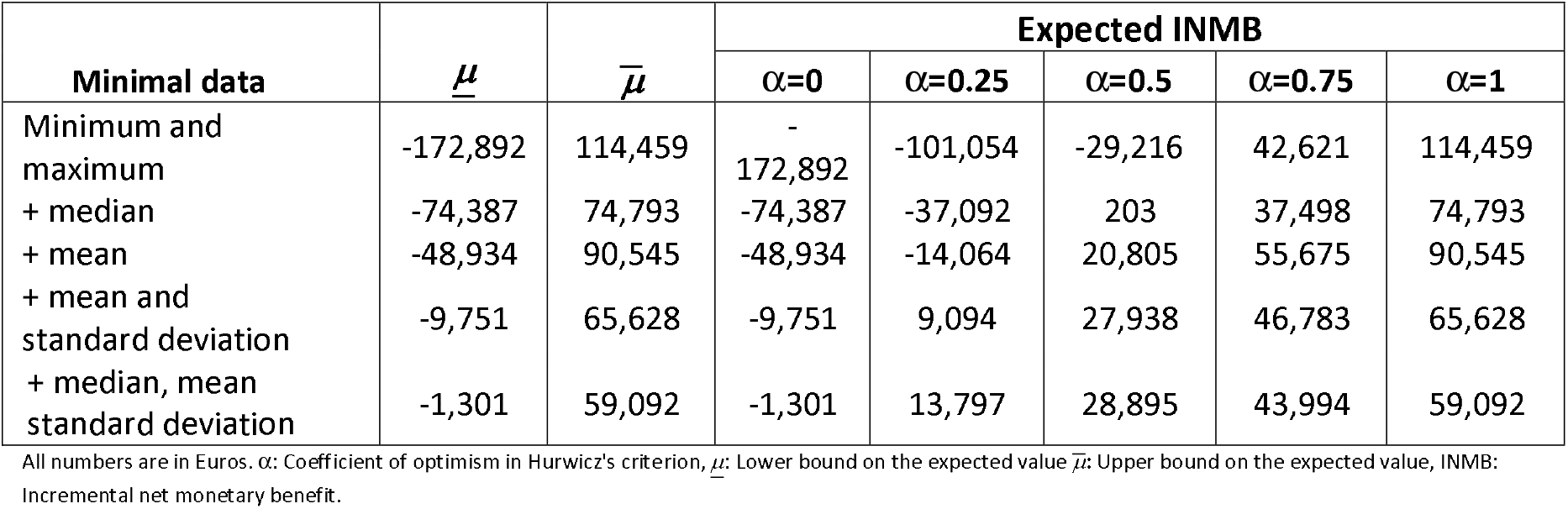
Expected incremental net monetary benefits of novel TAH using Hurwicz criterion under different minimal data.

## 5 DISCUSSION

This study introduces the probability bound analysis method for quantifying the effect of parameter uncertainty on decision-relevant outcomes in the context of an early assessment of novel health technologies characterized by data sparsity. This paper is also the first application of PBA that provides an alternative approach for modelling parameter uncertainty without necessitating the use of unverifiable assumptions as in the status quo approach (PSA). To assist practitioners, we provide step-by-step guidance and demonstrate an application of PBA to evaluate the cost-effectiveness of a novel total artificial heart.

### 5.1 Advantages of PBA

The PBA method is more attractive than the standard approach in the following ways. First, in PBA, parameter uncertainties are characterized by p-boxes that provide the maximum area of uncertainty (tightest bounds) containing the unknown CDF, given some data characterizing the CDF (e.g., summary statistics). The uncertainty propagation of p-boxes into a black-box model, using optimization, generates bounds that are guaranteed to enclose *all possible CDFs* of the outcomes of interest provided that the p-boxes of the parameters enclose their respective (unknown) CDFs. Second, if the lower and upper bounds of a CDF coincide for every element in the parameter support, then a p-box degenerates to a CDF: a situation where Monte Carlo simulation is the standard approach. Therefore, PBA is a generalization of approaches that imply a full characterization of parameters distributions (e.g., PSA and expert elicitation). Thirdly, when analysts have data only the minimum and maximum values, PBA allows them to encode their uncertainty using a p-box without needing to assume the likelihood of the values between these two bounds. If, in addition to the extreme points, the analyst has data on the mean but not the standard deviation, PBA allows them to construct a p-box. This is an improvement over the common practice to assume that the standard deviation is some fractions of the mean (e.g., 20%), which is an artificial, non-justifiable, and often being hand-waived, approach to represent uncertainty. Fifth, critical decision-relevant information from a PBA, as demonstrated by the first comparison in our case study, are the bounds on the plausible values of a model outcome. This information is particularly useful when the outcome represents a decisional criterion, such as the INMB in our case study. The resulting p-box that spans over both positive and negative INMB values suggests that there is substantial uncertainty in the INMB being positive as well as negative. The status-quo approach may yield less uncertainty in the INMB and potentially lead to an over-confidence in the INMB being positive. Lastly, PBA provides a way to systematically evaluate how our degree of optimism towards the best-case scenario in combination with the degree of uncertainty (represented by the data availability) affects the optimal decision or the cost-effectiveness of a new technology.

### 5.2 Challenges of PBA

Despite its utility, PBA is computationally intensive for the following reasons. First, implementing PBA requires an optimization step over the parameter space. The higher the desired level of accuracy is, the higher number of sub-intervals *n*_*i*_ (i.e., the finer the discretization of the parameter space) is needed. Second, an increase in the number of p-box parameters will lead to a higher-dimensional optimization problem. Third, the computational cost is further exacerbated if the model is “expensive” to evaluate for a given set of parameter values. Fourth, if, in addition to p-box parameters, some parameters are characterized by their precise CDFs, the optimization step is embedded in a Monte Carlo sampling loop (**S0** and **S8**); thereby increasing the number of optimizations by a factor of N (the total number of Monte Carlo samples). To decrease the computational cost, practitioners may opt to use more efficient optimization methods [41], fast-to-evaluate approximations of the original model or meta-models [42], parallelization of step S6 by distributing the optimization task across computing units (e.g., central processing unit cores) in combination with using high-performance computing [43], and a more efficient design-of-experiment for steps S3 and S4.[44] Nevertheless, we expect a higher computational burden since a PBA imposes fewer restrictions (i.e., we do not assume a functional form), leading to a larger region of uncertainty over which a model needs to be evaluated. With the rapid advancement of computing power and capacity, however, it is only a matter of time before the higher computational burden is no longer a constraint. When the first health economic models and PSA emerged in the 1980s, the models were computationally intensive for the computing power available at the time; however, today they no longer pose a significant computational challenge.

### 5.3 PBA in regulatory decisions and health technology assessment

Health care decision-makers often do not consider the effect of the underlying assumptions of methods that analysts employ in health technology assessment. This ignorance is problematic from both regulatory and reimbursement perspectives, as it may result in technologies with an unfavourable benefit-risk profile entering the market (regulatory decision) or in products that are not cost-effective being reimbursed (reimbursement decision). Compared to PSA, PBA requires fewer assumptions of the input parameters, i.e., on their distribution, but it gives the same answer as PSA if the information is abundant enough [45]. PBA is especially advantageous for the assessment of medical technologies, where the uncertainty in the decision-making on regulatory but also on reimbursement stage is very high, i.e., the area between the two probability bounds is very large. High uncertainty on efficacy and effectiveness have been observed in medical devices entering the market with much lower regulatory requirements than medicinal products [46]. Similarly, orphan drugs and advanced therapy medicinal products usually obtain market approval with very small clinical studies [47] or the use of surrogate and intermediate endpoints [48]. In such situations where information on uncertainty about the relevant endpoints is needed and is hardly available, PBA is superior to PSA. In contrast to PSA that reduces the uncertainty to a single (precise) CDF of a decision-relevant endpoint and a rather narrow range of possible values, PBA gives decision-makers a region of uncertainty that is likely very large for medical devices, orphan drugs, and advanced medicinal products for the aforementioned reasons. Within the PBA framework, the shortcoming in the evidence base, the resulting uncertainty, and the limitation in the analytical results used for aiding decision-makers are made explicit; thereby promoting responsible decision-making

As mentioned, the ISPOR-SMDM Modeling Good Research Practices Task Force recommends that when there is very little information on a parameter, expert opinion should be elicited. Should elicitation methods be used, the best practice recommends the use of a conservative approach where the uncertainty analysis incorporates an appropriately broad range of possible estimates elicited from each expert and pooled together. Expert opinion elicitation requires that experts express an opinion a number of summary measures to allow statistical fitting of a distribution. Nonetheless, in some cases, experts may struggle to perform such task, because what they are called to express a judgment on may be still greatly unknown to them. In such cases, PBA may be used as an alternative method to characterize uncertainty without requiring too much elicitation from the experts. For example, experts may express a judgment only on the minimum and maximum values of the parameters or their median, and then PBA could be performed using only these summary measures. The characterization of the uncertainty is also strictly related to the assessment of whether further research would be worthwhile.

Although the PBA has advantages to PSA, the choice of methods in health technology assessment is very much dependent on whether methods are included in the guidance documents of national HTA agencies, such as the English NICE, the German IQWiG, or HAS in France. However, PBA is not the standard in the assessment of medicinal products yet. For medical devices, HTA guidance is scarce, even when viewed internationally [49, 50]. Nevertheless, the current HTA environment is characterized by massive momentum where new methods and concepts are sought and explored to reduce the burden of the manufacturers to fulfil diverging country requirements. In this regard, the activities of the European Union on the Regulation on Health Technology Assessment [51] could open a window of opportunity to establish PBA in future European HTAs. In particular, an internationally coordinated methods document could help PBA achieve a breakthrough in the assessment of medical devices and other medical technologies, which are characterized by deep uncertainty and improve the methodological basis for decision-making in health care.

### 5.4 Limitations

Our study has limitations in the following context. First, we assume independence among the model parameters. To the extent of our knowledge, how to model dependencies among the parameters in the context of uncertainty propagation using PBA and black-box models is an open problem and warrants further study. Secondly, we do not prescribe rules on when one should adopt the PBA approach or other methods to characterize parameter uncertainty. Such decisions are problem-dependent and left to the analysts. For example, a parameter may be highly uncertain due to the lack of empirical data and/or previous knowledge and, at the same time, non-influential, i.e., the decision is not sensitive to variations in the parameter values. However, there are clear-cut situations, including those where only the minimum and maximum values or minimum, maximum, and mean values are available, as mentioned above. In principle, we recommend the use of PBA as a complementary approach to PSA, whenever computationally feasible, as we tend to underestimate uncertainty in general [52]. Thirdly, we do not conduct a formal error analysis and, instead, provide heuristics to choose the optimal number of sub-intervals. An analysis of the approximation error, i.e., determining its rate of convergence to zero or estimating its upper bound, is beyond the scope of the paper. Moreover, the existence of an upper bound for the error is problem-dependent because the metric to calculate the error (e.g., the relative difference in INMB estimates) depends on the decision-making problem. Lastly, we demonstrate only one usage of the results from PBA, i.e., how to identify the optimal intervention in decision analysis. Since a PBA generates bounds on the unknown CDF, the expected value is interval-valued instead of a single value. Therefore, a re-formulation of approaches that rely on a single expected value, including value of information analyses, is needed and is, however, beyond the scope of our study. A more comprehensive exploration of how to utilize the results of a PBA should be the focus of future studies.

## 6 CONCLUDING REMARKS

With steadily rising costs, it is increasingly important that only effective and cost-effective technologies are implemented in the healthcare system. Decision-makers at care provider and administrative levels are responsible and accountable for adopting innovative technologies at the early stages under great uncertainty. To improve the information basis for their decision-making, HTA agencies should continuously monitor the field and assess new methods for the assessment of health technologies. Approaches that have demonstrated methodological superiority should be integrated into the methodological guidelines. With the proposed parameter uncertainty quantification approach, we provide a tool that generalizes current methodologies for characterizing uncertainty in data and knowledge used to inform health economic models. PBA does not rely on unverifiable assumptions and will make decision-making more transparent, leading to better and more responsible decisions for patients and for a sustainable health care system.

## Data Availability

All data produced in the present work are contained in the manuscript

## A APPENDIX

### A.1 P-Box formulas

This section lists the p-box formulas for common situations of data availability where a modeler can identify and specify a combination of different summary statistics of and/or information on θ.

The first situation involves knowing the smallest (minimum) and largest (maximum) values of a parameter. For some parameters, one can infer the range from theoretical limits, such as zero to one for probability or utility parameters (if zero represents the lowest utility value). In some cases, a modeler may ask domain experts to specify a range from their knowledge about the quantity in question. In both cases, the task will set *a* and *b* such that the parameter lies in the interval (*I*)bounded by *a* and *b* : θ ∈ [a,b]: = I_*θ*_. The p-box 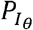 is given by:

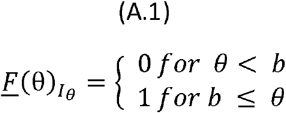

for LBF, and,

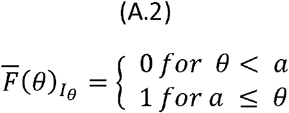

for UBF.

The inverse functions of the p-box 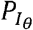 are given by:

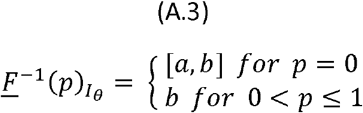

For LBF, and,

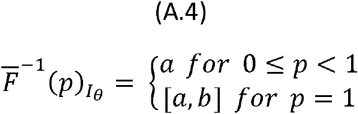

For UBF.

If, in addition to knowing *I*_*θ*_, the median *m* of θ is also known, then the p-box can be tighter. The p-box 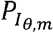 is given by:

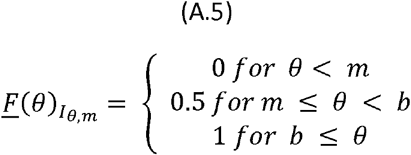

for LBF, and,

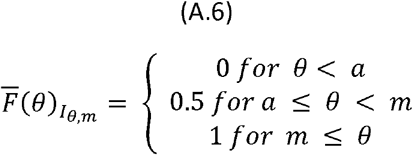

for UBF.

The inverse functions of the p-box 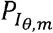 are given by:

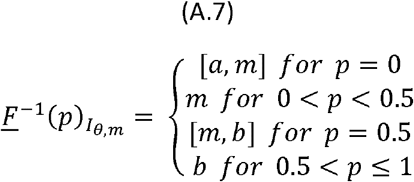

For LBF, and,

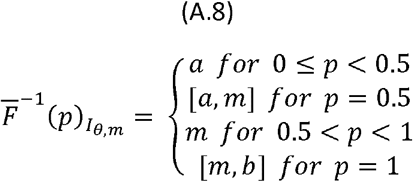

For UBF.

If, in addition to knowing *I*_*θ*_, the mean *µ* = *E* [*θ*]is also known, then the p-box 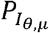, is given by:

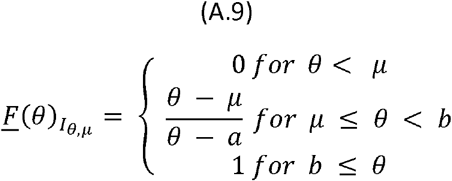

for LBF, and,

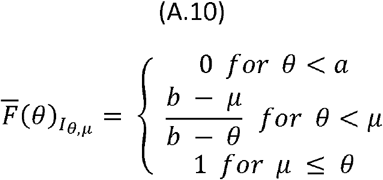

for UBF.

The inverse functions for the p-box 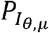, are given by:

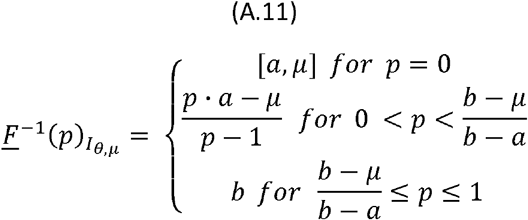

For LBF, and,

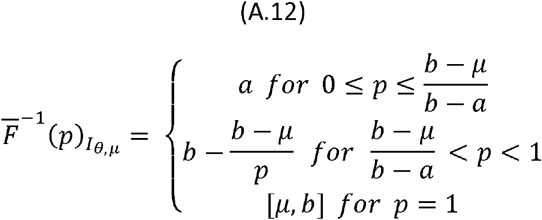

for UBF.

If, in addition to knowing *I*_*θ*_, and *µ*, we have data on the standard deviation *σ*, then the p-box 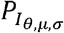, is given by:

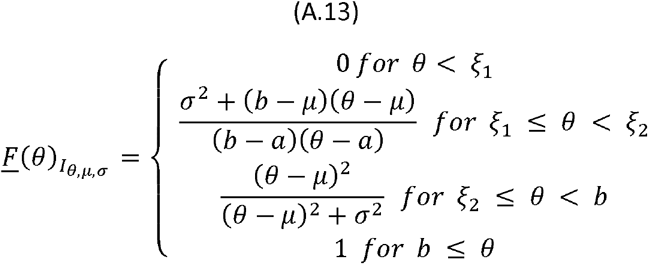

for LBF, and,

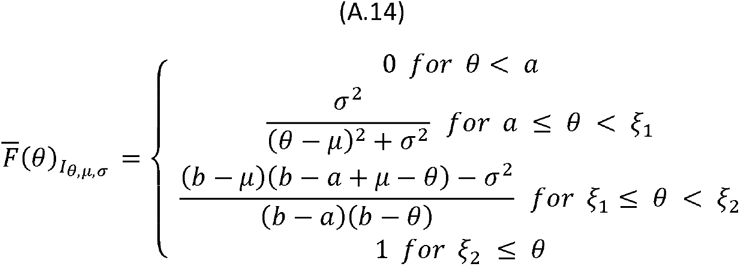

for UBF, where 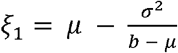 and 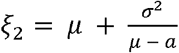.

The inverse functions for the p-box 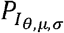, are given by:

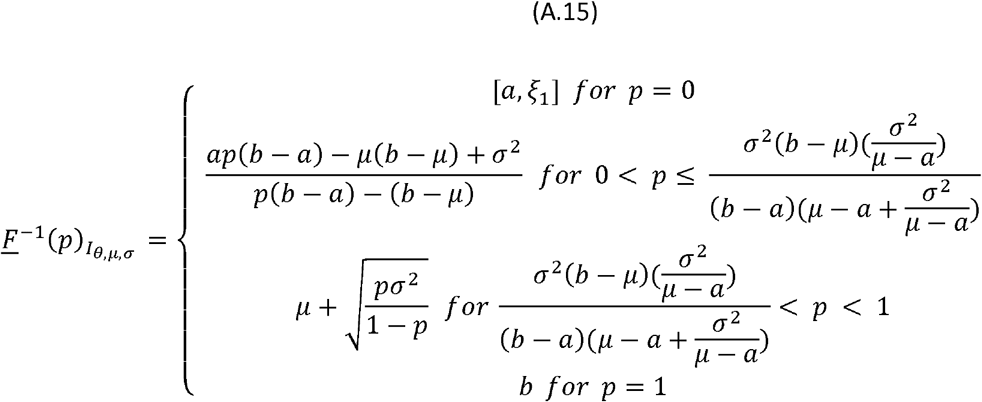

for LBF, and,

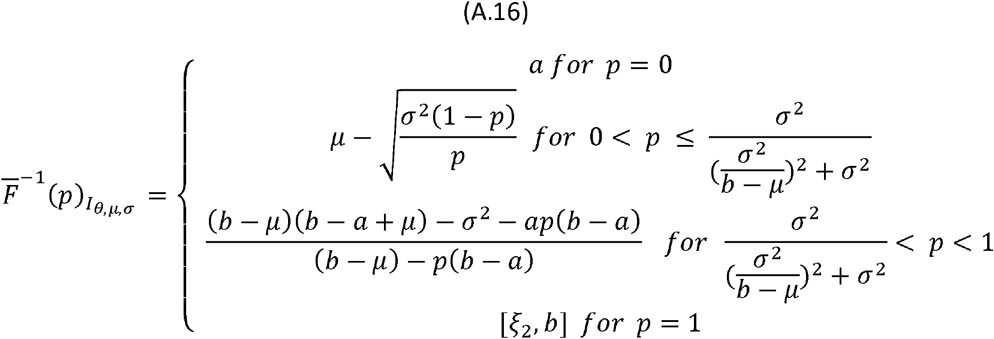

for UBF, where 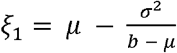 and 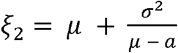.

In principle, as we have additional summary statistics on θ, or more information about the unknown *F*(*θ*), the p-box becomes tighter. In general, we can derive further cases by intersecting the p-boxes of different 𝒟s described above (termed as primitive p-boxes) by “tracing” the minimum (or maximum) of the intersection of the corresponding UBFs (or LBFs). For each *θ* value in the support, we compute the p-boxes for all 𝒟s, which are included in the intersection and whose formulas are known. Then, given a *θ* value, we find the minimum (maximum) value of the UBFs (LBFs) and set this value as the UBF (LBF) value of the intersected p-boxes. We then repeat the procedure for all *θ* values and trace values to produce the LBF and UBF. As an example, we can derive the p-box for median) by intersecting the p-box for 𝒟 = {*I*_*θ*,_ *µ*, *σ*} (where m is the median) by intersecting the p-box for 𝒟 = {*I*_*θ*,_*m*} (the top-right subplot of Figure 1) and the p-box for 𝒟 = {*I*_*θ*,_ *µ*, *σ*} (the bottom-center subplot of Figure 1). We can superimpose one plot over the other and apply the approach to trace the intersected p-box. More formally, for each 𝒟_*d*_ (where d indexes each combination of available data), the LBF and UBF are given by:

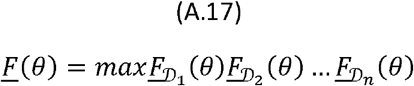

and

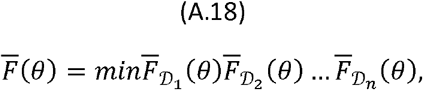

respectively.

### A.2 Supplementary tables

**Table A.2.** parameters for the Syncardia from secondary sources.

| Values | Mean (95% Credible Interval) | Source |
| --- | --- | --- |
| **Probability of remaining in the TAH state – Survival curve parameters** |  |  |
| Location parameter TAH | 5.1 | [53] |
| Shape parameter | 0.9 | [53] |
| **Probability of death while on support conditional on leaving the TAH state** |  |  |
| 1 month | 0.7 | [53] |
| 2 month | 0.5 | [53] |
| 3 month | 0.4 | [53] |
| > 3 months | 0.2 | [53] |
| **Complications while on support** |  |  |
| Strokes | 0.23 | [53] |
| Proportion of disabling strokes | 0.12 moderate-severe disability; 0.39 Severe | [53] |
| Major bleeding | 0.21 | [53] |
| Major Infection | 0.7 | [53] |
| Major device malfunction | 0.106 | [53] |
| **Survival after HTx** |  |  |
| mu parameter | -4.728 | [54] |
| Sigma parameter | 0.920 | [54] |
| Shape parameter | -29.795 | [54] |
| **Quality of Life** |  |  |
| Quality of life while on support | 0.64 | Assumed |
| Quality of life after HTx – without disabling stroke | 0.76 | [35] |
| Long term utility decrement with disabling stroke | -0.18 | [55] |
| **Costs** |  |  |
| Cost heart transplant procedure and first month in the hospital |  |  |
| **TAH cost** | € 100,000 | Expenses data from the Italian Ministry of health |
| TAH implant procedure + first month hospitalization | 156,625 | [35] |
| Monthly hospitalization cost after the first | Decreasing from 19, for the 2<sup>nd</sup> month to 1,744 in the 8<sup>th</sup> month | [35] |
| Proportion of TAH patients discharged | 0.11 | [53] |
| Average LOS before discharge (for those discharged) | 50 | [53] |
| Costs of Stroke | 5,587 | Italian DRGs 559; 14 and 15 (mean of the three values) |
| Cost of Major bleeding requiring re-operation | 4,321 | Italian DRG 174 |
| Cost of major infection | 7,148 | Italian DRG 576 |
| Cost of major device malfunction | 81,551 | Italian DRG 103 |

**Table A.3.** Parameters simulated for the novel TAH.

|  |  |  |
| --- | --- | --- |
| **Values** | **Mean (95% Credible Interval)** | **Parametric distribution used in PSA** |
| **Probability of remaining in the TAH state – Survival curve parameters** |  |  |
| Location parameter – novel TAH | 4.14 |  |
| Shape parameter | 0.9 |  |
| **Probability of death while on support conditional on leaving the TAH state** |  |  |
| 1 month | 0.61 (0.34; 0.84) | Beta |
| 2 month | 0.12 (0.003; 0.41) | Beta |
| 3 month | 0.14 (0.004; 0.43) | Beta |
| > 3 months | 0.13 |  |
| **Complications while on support** |  |  |
| **Strokes** | 0.21 |  |
| **Major bleeding** | 0.19 |  |
| **Major Infection** | 0.59 |  |
| **Major device malfunction** | 0.108 (0.03; 0.22) | Beta |
| **TAH cost** | € 200,000 |  |
| Proportion of TAH patients discharged | 0.73 |  |

## References

1. Sculpher, M., M. Drummond, and M. Buxton, The iterative use of economic evaluation as part of the process of health technology assessment. Journal of health services research & policy, 1997. 2(1): p. 26–30.

2. Mauskopf, J., et al., A strategy for collecting pharmacoeconomic data during phase II/III clinical trials. Pharmacoeconomics, 1996. 9(3): p. 264–277.

3. Grabowski, H. and C.D. Mullins, Pharmacy benefit management, cost-effectiveness analysis and drug formulary decisions. Social science & medicine, 1997. 45(4): p. 535–544.

4. Terrés, C.R., Pharmacoeconomic analysis in new drug development: a pragmatic approach to efficiency studies. Clinical research and regulatory affairs, 1998. 15(3-4): p. 209–223.

5. Grutters, J.P., et al., Problems and promises of health technologies: the role of early health economic modeling. International journal of health policy and management, 2019. 8(10): p. 575.

6. IJzerman, M.J., et al., Emerging use of early health technology assessment in medical product development: a scoping review of the literature. Pharmacoeconomics, 2017. 35(7): p. 727–740.

7. Ijzerman, M.J. and L.M. Steuten, Early assessment of medical technologies to inform product development and market access. Applied health economics and health policy, 2011. 9(5): p. 331–347.

8. Markiewicz, K., J.A. Van Til, and M.J. IJzerman, Medical devices early assessment methods: systematic literature review. Int J Technol Assess Health Care, 2014. 30(2): p. 137–146.

9. Hartz, S. and J. John, Contribution of economic evaluation to decision making in early phases of product development: a methodological and empirical review. International Journal of Technology Assessment in Health Care, 2008. 24(4): p. 465–472.

10. Miller, P.S., F.L. Andersson, and L. Kalra, Are cost benefits of anticoagulation for stroke prevention in atrial fibrillation underestimated? Stroke, 2005. 36(2): p. 360–366.

11. Drummond, M.F., Modeling in Early Stages of Technology Development: Is an Iterative Approach Needed?: Comment on” Problems and Promises of Health Technologies: The Role of Early Health Economic Modeling”. International Journal of Health Policy and Management, 2020. 9(6): p. 260.

12. McKenna, C. and K. Claxton, Addressing adoption and research design decisions simultaneously: the role of value of sample information analysis. Medical decision making, 2011. 31(6): p. 853–865.

13. Love-Koh, J., How Useful Are Early Economic Models?: Comment on” Problems and Promises of Health Technologies: The Role of Early Health Economic Modelling”. International Journal of Health Policy and Management, 2020. 9(5): p. 215.

14. Backhouse, M.E., et al., Early dialogue between the developers of new technologies and pricing and reimbursement agencies: a pilot study. Value in Health, 2011. 14(4): p. 608–615.

15. Briggs, A., Probabilistic analysis of cost-effectiveness models: statistical representation of parameter uncertainty. Value in Health, 2005. 8(1).

16. Bilcke, J., et al., Accounting for methodological, structural, and parameter uncertainty in decision-analytic models: a practical guide. Medical Decision Making, 2011. 31(4): p. 675–692.

17. Briggs, A.H., et al., Model parameter estimation and uncertainty analysis: a report of the ISPOR-SMDM Modeling Good Research Practices Task Force Working Group–6. Medical decision making, 2012. 32(5): p. 722–732.

18. O’Hagan, A., et al., Uncertain judgements: eliciting experts’ probabilities. 2006.

19. Iskandar, R., Probability bound analysis: A novel approach for quantifying parameter uncertainty in decision-analytic modeling and cost-effectiveness analysis. Statistics in Medicine, 2021. 40(29): p. 6501–6522.

20. Ferson, S. and L.R. Ginzburg, Different methods are needed to propagate ignorance and variability. Reliability Engineering & System Safety, 1996. 54(2-3): p. 133–144.

21. Massey Jr, F.J., The Kolmogorov-Smirnov test for goodness of fit. Journal of the American statistical Association, 1951. 46(253): p. 68–78.

22. Feller, W., An introduction to probability theory and its applications, vol 2. 2008: John Wiley & Sons.

23. Doubilet, P., et al., Probabilistic sensitivity analysis using Monte Carlo simulation: a practical approach. Medical decision making, 1985. 5(2): p. 157–177.

24. Ferson, S., et al., Constructing probability boxes and Dempster-Shafer structures. 2015, Sandia National Lab.(SNL-NM), Albuquerque, NM (United States).

25. Williamson, R.C. and T. Downs, Probabilistic arithmetic. I. Numerical methods for calculating convolutions and dependency bounds. International journal of approximate reasoning, 1990. 4(2): p. 89–158.

26. Iskandar, R., Probability bound analysis: A novel approach for quantifying parameter uncertainty in decision-analytic modeling and cost-effectiveness analysis. arXiv preprint 2011.10398, 2020.

27. Arabía, F.A., The Total Artificial Heart: Where Are We? Cardiology in Review, 2020. 28(6): p. 275–282.

28. Copeland, J.G., et al., Cardiac replacement with a total artificial heart as a bridge to transplantation. New England Journal of Medicine, 2004. 351(9): p. 859–867.

29. Netuka, I., et al., Fully magnetically levitated left ventricular assist system for treating advanced HF: a multicenter study. Journal of the American College of Cardiology, 2015. 66(23): p. 2579–2589.

30. Strueber, M., et al., Results of the post-market registry to evaluate the HeartWare left ventricular assist system (ReVOLVE). The Journal of Heart and Lung Transplantation, 2014. 33(5): p. 486–491.

31. de By, T.M., et al., The European Registry for Patients with Mechanical Circulatory Support (EUROMACS) of the European Association for Cardio-Thoracic Surgery (EACTS): second report. European journal of cardio-thoracic surgery, 2018. 53(2): p. 309–316.

32. Molina, E.J., et al., The Society of Thoracic Surgeons Intermacs 2020 Annual Report. The Annals of thoracic surgery, 2021. 111(3): p. 778–792.

33. Schmier, J.K., et al., A systematic review of cost-effectiveness analyses of left ventricular assist devices: issues and challenges. Applied health economics and health policy, 2019. 17(1): p. 35–46.

34. Federici, C. and L. Pecchia, Exploring the misalignment on the value of further research between payers and manufacturers. A case study on a novel total artificial heart. 2021.

35. Sharples, L., et al., Evaluation of the ventricular assist device programme in the UK. HEALTH TECHNOLOGY ASSESSMENT-SOUTHAMPTON-, 2006. 10(48).

36. Computing, R., R: A language and environment for statistical computing.

37. Gablonsky, J.M. and C.T. Kelley, A locally-biased form of the DIRECT algorithm. Journal of Global Optimization, 2001. 21(1): p. 27–37.

38. Johnson, S.G., The NLopt nonlinear-optimization package. 2014.

39. Ypma, J., Introduction to nloptr: an R interface to NLopt. R Package, 2014. 2.

40. Ferson, S., L. Ginzburg, and R. Akçakaya, Whereof one cannot speak: when input distributions are unknown. Risk Analysis, 1996.

41. Deng, W., et al., An improved self-adaptive differential evolution algorithm and its application. Chemometrics and intelligent laboratory systems, 2013. 128: p. 66–76.

42. Ellis, A.G., et al., Active learning for efficiently training emulators of computationally expensive mathematical models. Statistics in Medicine, 2020. 39(25): p. 3521–3548.

43. Kurgalin, S. and S. Borzunov, A practical approach to high-performance computing. Vol. 206. 2019: Springer.

44. Schöbi, R. and B. Sudret, Uncertainty propagation of p-boxes using sparse polynomial chaos expansions. Journal of Computational Physics, 2017. 339: p. 307–327.

45. Zio, E. and N. Pedroni, Literature review of methods for representing uncertainty. 2013.

46. Shatrov, K.B. C.R., In the midst of a regulatory turmoil: Is the new European medical device regulation likely to achieve its main goals? 2021.

47. Pontes, C., et al., Evidence supporting regulatory-decision making on orphan medicinal products authorisation in Europe: methodological uncertainties. Orphanet journal of rare diseases, 2018. 13(1): p. 1–15.

48. Grigore, B., et al., Surrogate endpoints in health technology assessment: an international review of methodological guidelines. Pharmacoeconomics, 2020: p. 1–16.

49. Ciani, O., et al., Health technology assessment of medical devices: a survey of non-European union agencies. International journal of technology assessment in health care, 2015. 31(3): p. 154–165.

50. Fuchs, S., et al., HTA of medical devices: challenges and ideas for the future from a European perspective. Health Policy, 2017. 121(3): p 215–229.

51. European Commission, D.-G.f.H.a.F.S., Proposal for a Regulation of the European Parliament and of the Council on Health Technology Assessment and Amending Directive 2011/24/EU, SANTE, Editor. 2018.

52. Savage, S.L. and H.M. Markowitz, The flaw of averages: Why we underestimate risk in the face of uncertainty. 2009: John Wiley & Sons.

53. Arabía, F.A., et al., Interagency registry for mechanically assisted circulatory support report on the total artificial heart. The Journal of Heart and Lung Transplantation, 2018. 37(11): p. 1304–1312.

54. David, C.-H., et al., A heart transplant after total artificial heart support: initial and long-term results. European Journal of Cardio-Thoracic Surgery, 2020. 58(6): p. 1175–1181.

55. Luengo-Fernandez, R., et al., Quality of life after TIA and stroke: ten-year results of the Oxford Vascular Study. Neurology, 2013. 81(18): p. 1588–1595.

